# Dynamics of SARS-CoV-2-specific antibodies during and after COVID19: Lessons from a biobank in Argentina

**DOI:** 10.1101/2021.01.31.21250167

**Authors:** Yésica Longueira, María Laura Polo, InViV working group, Gabriela Turk, Natalia Laufer, Biobanco de Enfermedades Infecciosas Colección COVID19 working group

## Abstract

**Background:** Biobanks are instrumental for accelerating research. Early in SARS-CoV-2 pandemic, the Argentinean Biobank of Infectious Diseases (BBEI) initiated the COVID19 collection and started its characterization.

**Methods:** Blood samples from subjects with confirmed SARS-CoV-2 infection either admitted to health institutions or outpatients, were enrolled. Highly exposed seronegative individuals, were also enrolled. Longitudinal samples were obtained in a subset of donors, including persons who donated plasma for therapeutic purposes (plasma donors). SARS-CoV-2-specific IgM and IgG levels, IgG titers and IgG viral neutralization capacity were determined.

**Findings:** Out of 825 donors, 57.1% were females and median age was 41 years (IQR 32-53 years). Donors were segregated as acute or convalescent donors, and mild versus moderate/severe disease donors. Seventy-eight percent showed seroconversion to SARS-CoV-2 specific antibodies. Specific IgM and IgG showed comparable positivity rates in acute donors. IgM detectability rate declined in convalescent donors while IgG detectability remained elevated in early (74,8%) and late (83%) convalescent donors. Among donors with follow-up samples, IgG levels seemed to decline more rapidly in plasma donors. IgG levels were higher with age, disease severity, number of symptoms, and was more durable in moderate/severe disease donors. Levels and titers of anti-spike/RBD IgG strongly correlated with neutralization activity against WT virus.

**Interpretation:** The BBEI-COVID19 collection served a dual role in this SARS-CoV-2 global crisis. First, it feed researchers and developers transferring samples and data to fuel research projects. Second, it generated highly needed local data to understand and frame the regional dynamics of the *infection*.

**Funding:** This work was supported by a grant from the Agencia Nacional de Promoción de la Investigación, el Desarrollo Tecnológico y la Innovación (Agencia I+D+i) from Argentina through an extraordinary funding opportunity to improve the national response to COVID19 (Proyecto COVID N° 11, IP 285).

## INTRODUCTION

In December 2019, a cluster of cases of atypical interstitial pneumonia caused by an unknown agent was reported in China [1]. Afterwards, it was described that this new disease (termed COVID-19) was caused by a novel human coronavirus which was isolated, characterized and named Severe Acute Respiratory Syndrome Coronavirus 2 (SARS-CoV-2) [2]. Since then, the number of global cases has increased rapidly, with the WHO declaring COVID-19 a pandemic in March 2020. By the beginning of January 2021, more than 90 million cases have been confirmed worldwide, with more than 2.1 million associated deaths [3]. In Argentina, the first case was confirmed on March 3^rd^, 2020 and was a 43-year-old male returning from a trip around Spain and Italy. This report was followed by other imported cases and soon local circulation was established. The number of cases has reached 1.86 million (including near 46800 deaths) by January 24^th^, 2021 [4].

SARS-CoV-2 can be transmitted from human to human by respiratory droplets and aerosols, and close contact with infected people and contaminated objects. The infection can be symptomatic or asymptomatic. In most cases, symptoms appear within 48-72 hs after exposure and may include fever, cough, runny nose, odynophagia, headache, asthenia, myalgia, anosmia, ageusia, skin manifestations among others [5]. Although most subjects recover after experiencing a mild disease, a minority of individuals progress to a severe disease with symptoms and signs associated with viral pneumonia and pulmonary involvement, which may lead to the need of mechanical ventilation and death. Less frequently, neurological manifestations may present [6]. People with comorbidities such as diabetes, cardiovascular disease, kidney disease, hypertension, obesity, and chronic obstructive pulmonary disease are overrepresented among those with severe COVID-19 and those who died. Indeed, the fatality rate is particularly high in older patients, in whom comorbidities are common [7].

The COVID-19 pandemic brought a great challenge to society and, more specifically, to the health and scientific systems. Social containment measures have been adopted worldwide to stop virus dissemination. As it occurred in most countries around the globe, the Argentinean health-care system quickly adapted to cope with an overwhelming number of acutely ill patients, and the scientific community redirected their research to provide responses to the emergency, guided by the Ministry of Science. Although tremendous advances have been made, our understanding regarding the dynamics of the disease is not complete, slowing the processes of developing proper diagnostic algorithms, efficacious treatments, and preventive vaccines.

We have established the first national Biobank of Infectious Diseases in Argentina in 2017 (BBEI, *Biobanco de Enfermedades Infecciosas*). A biobank is a key tool in biomedical research, connecting basic and translational sciences. The proper and secure storage of large amounts of human biological samples from patients with specific conditions or healthy donors allows exploration and discovery of markers for pathological conditions, as well as identification and validation of new therapies [8]. At the same time, a biobank guarantees adherence to ethical and legal requirements in order to protect citizen rights [9]. Upon SARS-CoV-2 emergence, the BBEI rapidly initiated the collection of blood samples from confirmed or highly suspected COVID-19 subjects. The aim of the work is not only to create the sample collection but also to perform an in-depth clinical, immune and genetic characterization of our population.

This will allow to fuel research projects within the country by transferring samples and their clinical data but also to foster research by contributing additional laboratory data associated with these samples. Here, we present demographic, diagnostic, clinical and humoral response data of the initial 825 enrolled individuals. While the majority of donors with SARS-CoV-2 infection confirmed by molecular diagnostic seroconverted, a small proportion remained IgM and IgG negative. SARS-CoV-2 IgG response could be detected even at 5 months following symptoms onset with signs of waning by that time, particularly in the mild disease group. Finally, a steeper decay in IgG levels was observed in recovered participants that donated plasma for therapeutic purposes.

## MATERIALS AND METHODS

### BBEI COVID-19 cohort

The BBEI receive blood donations from subjects with confirmed SARS-CoV-2 infection by Real Time PCR or antibody testing. Donors can be inpatient, admitted to health institutions or outpatient individuals. They must provide written consent for the donation and a case report form (CRF) with clinical and demographic data is completed by the researchers. Additionally, highly exposed SARS-CoV-2 negative contacts (ES), defined as: persons who lived together with confirmed cases while they were symptomatic, but presented no evidence of infection, were enrolled [10]. The SARS CoV-2 collection within the BBEI was reviewed and approved by the institutional review board of the non-for-profit research organization Fundación Huésped (*Comité de Bioética Humana, Fundación Huésped, Buenos Aires, Argentina*)

### Data and sample collection

After signing the consent, 30 ml of whole blood were collected from donors in EDTA containing tubes (BD Vacutainer), and 10 ml in tubes containing no anticoagulants (SST tubes, BD Vacutainer). Following this, donors provided information regarding: gender, age, place of residence, whether they acquired the infection in Argentina or overseas, date of symptoms onset, date of diagnostics, comorbidities, treatments and complications; to complete the CRF. Among symptoms, donors were asked if they have experienced fever, headache, cough, expectoration, runny nose, dyspnea, odynophagia, asthenia, myalgia, anosmia, ageusia, anorexia, nausea, vomiting, diarrhea and/or skin manifestations. Every other relevant clinical data such as laboratory and image findings, hypoxemia, need of oxygen therapy and ventilation, were recorded.

### Sample processing

Blood samples were processed within 4 hours from withdrawal. Tubes were centrifuged at 2500 rpm for 10 minutes. Serum was separated from tubes without anticoagulants, aliquoted and stored at −80°C. Plasma was separated from EDTA-containing tubes, aliquoted and stored at −80°C. Then, blood was diluted and peripheral blood mononuclear cells (PBMCs) were isolated by Ficoll-Hypaque density gradient centrifugation (GE Healthcare, Sweden). Two pellets of 1 million cells were stored at −80°C for nucleic acid extraction and the remaining cells were preserved in a solution of fetal bovine serum (FBS, Sigma-Aldrich, USA) supplemented with 10% DMSO (Sigma-Aldrich) and stored in liquid nitrogen.

### Antibody assessment

The presence of SARS-CoV-2-specific IgM and IgG antibodies were evaluated in plasma samples from all donors enrolled in the biobank by ELISA using the COVIDAR kit. This kit was developed by Argentinean researchers from CONICET, Fundación Instituto Leloir and UNSAM together with Laboratorio Lemos S.R.L. The validation process determined that sensitivity to detect specific IgG raises to 90% after 3 weeks of symptoms onset while specificity is 100% [11]. Briefly, samples were loaded in wells pre-coated with recombinant SARS-CoV-2 Spike protein and RBD.

After incubation, wells were washed and HRP-conjugated anti-human IgG (or IgM) was added. Finally, the plates were developed using TMB substrate. Cut-off was calculated as the mean of the negative controls + 0.2 (IgG) or + 0.3 (IgM). Normalised optical density (NOD) values were calculated by subtracting the cut-off value to each donor sample OD value, and the resulting value was divided by the mean positive control OD value. In a selected subset of donors, SARS-CoV-2-specific IgG was titrated by making 2-fold serial dilutions of plasma.

Additionally, a subset of selected samples were also evaluated using an in-house indirect immunofluorescence assay (IFA) and by the commercially available kit Abbott Alinity i SARS-CoV-2 IgG. IFA was carried out by the InViV working group. Briefly, Vero Clon76 cells (ATCC CRL-587) were inoculated with SARS-CoV-2 strain (hCoV-19/Argentina/PAIS-G0001/2020, GISAID, ID: EPI_ISL_499083) at a multiplicity of infection (moi)= 0,1. Twenty-four hours post-infection, cells were harvested, seeded into slides (10.000 cells/well) and then fixed using cold (4°C) acetone during 30 min. Slides seeded with uninfected cells were also prepared as negative controls. Donor sera were diluted 1:5 in 1X PBS and added onto slides containing infected or uninfected cells. Slides were incubated at 37°C for 30min, washed, and then stained with FITC-labelled anti-human IgG for 30 min. After incubation, slides were washed and observed in a fluorescence microscope (Olympus Motorized Inverted Research, Model IX81, Imaging Software: Cell M). Samples were considered positive for SARS-COV-2 IgG antibodies when the specific apple-green fluorescence was located in the cytoplasm or on the plasma membrane in approximately 80% of the cells and no fluorescence staining was observed in the corresponding uninfected control.

Abbott Alinity i SARS-CoV-2 IgG kit, which captures SARS-CoV-2 N-specific antibodies, was used following manufacture’s instructions.

### Virus neutralization

Vero-E6 cells were maintained in DMEM medium (Sigma-Aldrich) plus 2 mM L-glutamine (Sigma-Aldrich), 100 U/ml penicillin (Sigma-Aldrich), 100 μg/ml streptomycin (Sigma-Aldrich) and 10% FBS (Gibco BRL, USA). SARS-CoV-2 strain (hCoV-19/Argentina/PAIS-G0001/2020, GISAID Accession ID: EPI_ISL_499083) was kindly provided by Dr. Sandra Gallego (InViV working group). Serial 2-fold dilutions of decomplemented plasma were incubated with 200 plaque-forming units (PFU) of SARS-CoV-2 for 1 h at room temperature, in triplicates. Then, mixtures were added to 80% confluent Vero-E6 cell monolayers in 96-well plates and incubated at 37⍰°C for 1 h. Then, cells were washed and culture medium with 2% FBS was added. After 72 h, plates were fixed with 4% paraformaldehyde for 20 min at room temperature and stained using a 0.5% crystal violet dye solution in acetone and methanol. Neutralization titer was calculated as the inverse of the highest plasma dilution that showed an 80% cytopathic effect inhibition.

### Data analysis

All data (clinical, demographic and laboratory data) associated with each donor was kept at the Noraybanks software database (Noraybio, Spain). Upon admission to the biobank, each donor was provided with a code (de-identification) and kept anonymous for subsequent processes. Statistical analyses were performed using GraphPad Prism 7 (GraphPad Software), InfoStat [12], R project (R Foundation for Statistical Computing, Vienna, Austria) and SPSS software v.19.0 (SPSS Corp., Armonk, NY, USA). Antibody titer fluctuations after plasma donation were analyzed by using the lmer package. All tests were considered statistically significant when the p-values were <0.05.

## RESULTS

### Cohort description

Administrative tasks to start the COVID-19 collection within the BBEI began on March 27^th^, 2020. From April 9^th^to October 9^th^, samples from 825 donors were enrolled, at a rate of 6.65 samples per working day. Moreover, longitudinal samples were obtained from 37 donors. Out of these initial 825 donors, 5596 vials of plasma, 2287 vials of serum, 1616 vials of cell pellets and 4347 vials of cryopreserved PBMCs were generated and this material became available to those researchers who might request it. Overall, 57.1% donors were females (n=471) and median age was 41 years (IQR 32-53 years); 6.6% were between 16 and 25 years old, and 16.8% were older than 60. Within the first donations, imported cases were overrepresented; but the local/imported ratio was rapidly reverted as regional circulation increased over the weeks. Donors were segregated as acute donors (those whose samples were obtained within 15 days from symptom onset), early convalescent donors (those whose samples were obtained within 60 days from symptom onset) and late convalescent donors (those whose samples were obtained later than 60 days from symptom onset). In turn, donors included within these groups were segregated into those with mild or moderate/severe disease. The latter was defined by the presence of related complication such as pneumonia, hypoxemia or need of oxygen. Thus, six groups were defined: acute severe (AS, N=84), acute mild (AM, N=61), early convalescent severe (ECS, N=45), early convalescent mild (ECM, N=408), late convalescent severe (LCS, N=17), late convalescent mild (LCM, N=120) (Table 1, Figure S1A). We could also define a seventh group composed of highly exposed SARS-CoV-2 seronegative contacts (ES, N=45), identified as persons who lived together with confirmed COVID-19 cases while they were symptomatic, but presented no evidence of infection themselves (no symptoms and negative SARS-CoV-2 serology after 21 days of exposure). Since it is known that a proportion of recovered subjects from SARS-CoV-2 infection do not seroconvert, it is worth noting here that it cannot be categorically excluded that infection had indeed occurred in ES. Moreover, it cannot be affirmed that they are resistant to the infection as the possibility that they acquired the infection after sample donation to BBEI cannot be excluded. Finally, 45 donors could not be segregated into any of these groups so they were excluded from further analysis. This group included donors who resulted negative in antibody testing, had no record of molecular diagnostic and had no history of extremely close contact with a confirmed COVID-19 (Table 1).

**Table 1:** Subject characteristics.

| Groups:<br>Characteristics: | AS | AM | ECS | ECM | LCS | LCM | ES | P |
| --- | --- | --- | --- | --- | --- | --- | --- | --- |
|  | N=84 (10.2%) | N=61 (7.4%) | N=45 (5.5%) | N= 408 (49.5%) | N= 17 (2.1%) | N= 120 (14.5) | N=45 (5.5%) |  |
| Age (years) Median (IQR) | 57.5 (47.5-67.2) | 36.5 (29.5-48.5) | 49 (39.25-57.5) | 37 (31-47.7) | 57 (50.5-63) | 36 (29-49) | 37 (31-51) | 0.000 |
| Female sex (N, %) | 33 (39.3%) | 29(47.5%) | 19(42.2%) | 252(61.8%) | 9(52.9%) | 80(66.7%) | 23(51.1%) | 0.001 |
| Days since symptoms onset<br>Median (IQR) | 10 (7-13) | 8 (5-12) | 36 (21-48) | 38 (28-45) | 72 (64.5-88) | 80 (68-96) | 41 (24-66) | <0.0001 |
| Comorbidities (N, %) |  |  |  |  |  |  |  |  |
| HTN | 32 (38.1%) | 5 (8.2%) | 10 (22.2%) | 30 (7.4%) | 4 (23.5%) | 6(5%) | 4 (8.9%) | 0.000 |
| DBT | 10(19.2%) | 2(5%) | 5(13.9%) | 7 (2.6%) | 0 (0.0%) | 1(1.2%) | 3(7.7%) | 0.000 |
| Obesity | 19 (22.6%) | 5(8.2%) | 6(13.3%) | 19(4.7%) | 2 (11.8%) | 7(5.9%) | 2(4.4%) | 0.002 |
| DLP | 0 (0.0%) | 0(0.0%) | 2 (4.4%) | 3 (0.7%) | 0 (0.0%) | 0 (0.0%) | 1 (2.2%) | 0.141 |
| ASTHMA | 8 (9.5%) | 2 (3.3%) | 4(8.9%) | 12(2.9%) | 2(11.8%) | 3 (2.5%) | 0 (0.0%) | 0.10 |
| HIV infection | 0 (0%) | 1 (1.6%) | 1(2.2%) | 7 (1.7%) | 2 (11.8%) | 5 (4.2%) | 0 (0.0%) | 0.01 |
| COVID TREATMENT (N, %) | 16 (20%) | 1(1.7%) | 16(37.2%) | 8 (2%) | 6 (40%) | 1 (0.9%) | 0 (0.0%) | 0.000 |
| POSITIVE SARS CoV2 PCR<br>(N, %) | 84 (100.0%) | 61 (100.0%) | 40 (88.9%) | 356 (86.7%) | 16 (94.1%) | 63(52.5%) | 0 (0.0%) | 0.000 |
| IgG anti SARSCoV2 + (N, %) | 59 (70.2%) | 22 (36.1%) | 43 (95.6%) | 292 (71.6%) | 17(100.0%) | 96(80.0%) | 0 (0.0%) | 0.000 |
| IgM anti SARSCoV2 + (N, %) | 57 (67.9%) | 23 (37.7%) | 37 (82.2%) | 173 (42.4%) | 9 (52.9%) | 45 (37.5%) | 0(0.0%) | 0.000 |

AS: acute severe, AM: acute mild, ECS: early convalescent severe, ECM: early convalescent mild, LCS: late convalescent severe, LCM: late convalescent mild, ES: highly exposed SARS-CoV-2 seronegative contacts, HTN: arterial hypertension, DBT: diabetes, DLP: dyslipidemia.

Seventy-five percent of donors had confirmed SARS-CoV-2 infection by molecular diagnosis (PCR). For donors who have not been tested by PCR (because they didn’t accomplished for criteria of suspected case at the time of diagnosis), infection was confirmed by serology. Most frequent comorbidities included arterial hypertension, diabetes, obesity, dyslipidemia, asthma and HIV infection. Comorbidities were overrepresented in donors who were experiencing or had experienced severe disease. Also, 48 donors received treatment, mostly consisting in azithromycin or plasma from recovered subjects.

### Dynamics of antibody responses

In order to evaluate the levels of SARS-CoV-2-specific IgG and IgM responses across all groups, these antibodies were qualitatively measured by ELISA. Normalized ODs (NOD) were calculated in order to compare data from different runs. Out of 825 donors, 579 (78,7% of the cohort excluding ES) showed SARS-CoV-2 specific antibodies, either IgM, IgG or both. Also, we identified donors with positive molecular diagnostics of SARS-CoV-2 infection (positive PCR, N=620) and no detectable antibody levels (N=152/620; 24,5%). Within this group, 34 were sampled during the very first days (2-7 days) after symptom onset so it is likely that sampling occurred too early to detect specific antibodies. Figure 1 shows the levels of IgM and IgG specific responses following symptoms onset. SARS-CoV-2-specific IgM was detected within the first days following symptoms onset; 52.85% of acutely infected donors had detectable plasma IgM (Figure 1A). IgM NOD started to increase between day 14 and 21 and then it showed a downward curve with a detectability rate of 48.45% between days 14 and 60 post-symptom onset. However, it is worth noting that IgM remained detectable in a significant proportion of donors (37.78%) between day 60 and 105 after symptom onset. IgM was undetectable in samples obtained after day 106 following symptoms onset. SARS-CoV-2-specific IgG showed an ascendant slope within the first two weeks from symptom onset, reaching a plateau between days 14 and 21, and was detectable in samples obtained up to 154 days post-symptom onset (Figure 1B). Positivity rate was 54.3% within acutely ill donors, a proportion comparable to that of IgM. The proportion of IgG detectability remained elevated in early and late convalescent donors (74,8% and 83,0%, respectively). Figures 1C and 1D show log_10_ transformation of IgM and IgG NOD respectively. This transformation eliminates negative responses. Again, it can be observed that IgM response climbed early and showed a slightly descending curve becoming undetectable in donors whose samples were obtained at very late convalescent stages. On the other hand, IgG could be detected in samples obtained up to 120 days following symptom onset and then began a decay phase suggesting that IgG levels might decrease over time following this period. However, it must be noted that data from late time-points after symptom onset are represented by a low number of donors so these conclusions should be further confirmed by increasing sample size.

**Figure 1:**
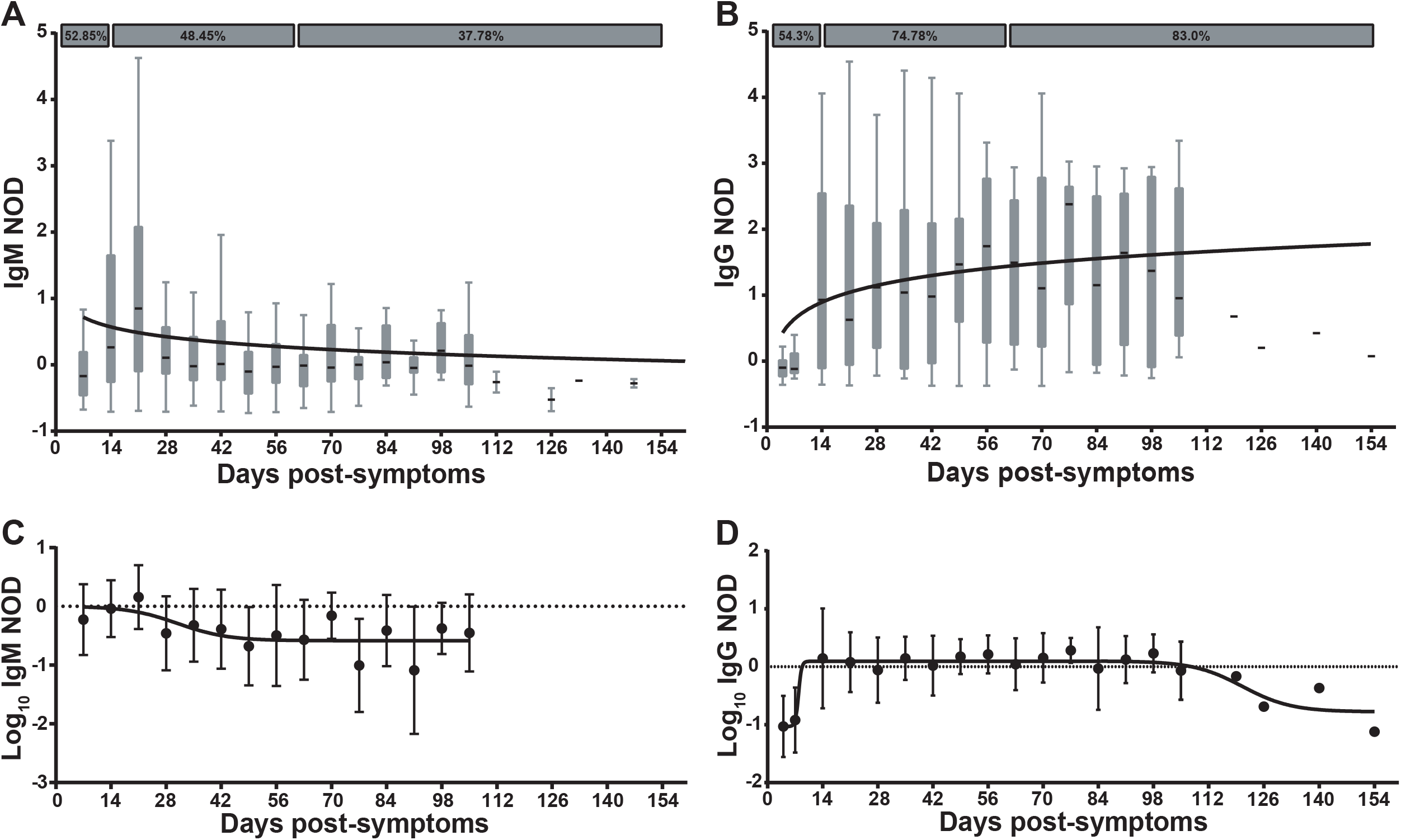
Dynamics of anti-SARS-CoV-2 IgM and IgG antibodies. **A)** Normalized optical density (NOD) measures for IgM, and **B)** IgG antibodies, were plotted by days post-symptoms onset. Positivity rates for acute, as well as early and late convalescent infection are indicated in both cases. **C)** Log10 of NOD values for IgM, and **D)** IgG antibodies were also obtained. Median and 25th and 75th percentiles are shown in A and B, while mean and standard errors are indicated in C and D. Longitudinal data was modeled by using a semi-log (A, B) and a sigmoidal 4PL non-linear regression model (C, D); best fitted curves are depicted in the plots.

Globally, no differences were observed in SARS-specific IgM or IgG responses between genders (not shown). SARS-specific IgM or IgG positive rates across groups is shown in Figure S1B. IgM and IgG rates are similar in the acute group. IgG rates were highest in convalescent donors (both early and late) while IgM was lowest in late convalescent. Of note, within each group (acute, early convalescent or late convalescent), higher positivity rates were observed in donors with severe/moderate disease compared to the mild disease group (donors who received plasma treatment were excluded from all analyses). When analyzing detectable IgG responses, older donors had higher levels of SARS-CoV-2-specific IgG (Figure 2A). In turn, IgG NOD was higher in donors with moderate/severe versus mild disease (Figure 2B, p<0.0001). It also increased with an increasing number of symptoms, being statistically higher in donors with ten or more symptoms compared with asymptomatic donors (p=0.0002) or donors reporting few symptoms (p=0.0078) (Figure 2C). Also, the probability of a positive IgG result increased as the number of symptoms augmented (p<0.0001, Figure 2D).

**Figure 2:**
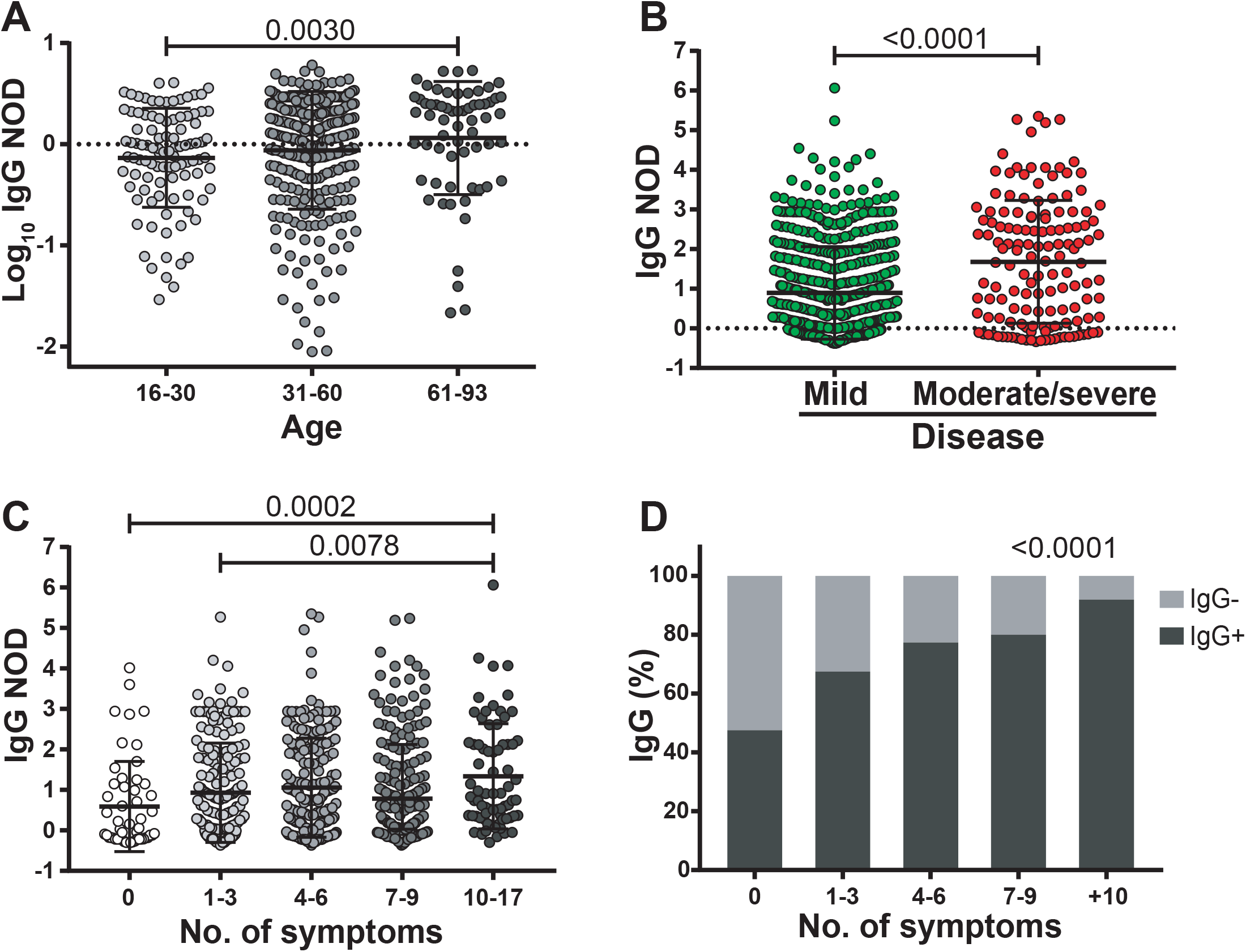
Characterization of IgG response in COVID-19 patients. **A)** Log10 of normalized optical density (NOD) IgG values from patients within separate age groups. **B)** NOD IgG levels from individuals with mild or moderate/severe disease. **C)** NOD IgG values from individuals displaying different number of symptoms. **D)** Positivity rate for IgG antibodies versus number of symptoms. Individual values, mean and standard deviation are shown. Statistical comparisons were made by using Kruskal-Wallis test followed by Dunn’s post-test (A, D), two-sided Mann-Whitney test (B) and chi squared test for trend (D).

### IgG titers and neutralization capacity

SARS-CoV-2-specific IgG titers were measured in a subset of donors (N=119). Logarithmic IgG titers showed a strong direct correlation with Log_10_ (IgG NOD) (Linear regression R^2^=0,9486, Spearman’s correlation r=0.9762, p=0,0004), indicating that Log_10_(IgG NOD) stands as a good surrogate for IgG response quantitation (Figure 3A). Analysis of IgG titers along time (in samples from different donors) revealed that this parameter, albeit showing a great heterogeneity among donors, remained rather stable during convalescent stage and up to 105 days after symptom onset (Figure 3B). Donors who experienced a severe disease tended to show elevated and stable antibody titers after recovery compared to the mild disease group, which in turn showed slight decrease in antibody titers over time. Indeed, statistical modeling of data showed that antibody titers remained steadily high long after the onset of symptoms in moderate/severe patients, while in the mild disease group, a slightly decrease in antibody titer was observed, confirming both groups behaved differently (p=0.0227). Moreover, when IgG titers measured after 15 days from symptom onset were pooled both in donors with mild or moderate/severe disease, the latter group showed statistically higher titers (p<0.0001; Figure 3C). Also, IgG titers were measured in asymptomatic subjects. This group showed similar titers to the mild disease group and statistically lower titers than the severe disease group (p=0.0007, Figure 3C).

**Figure 3:**
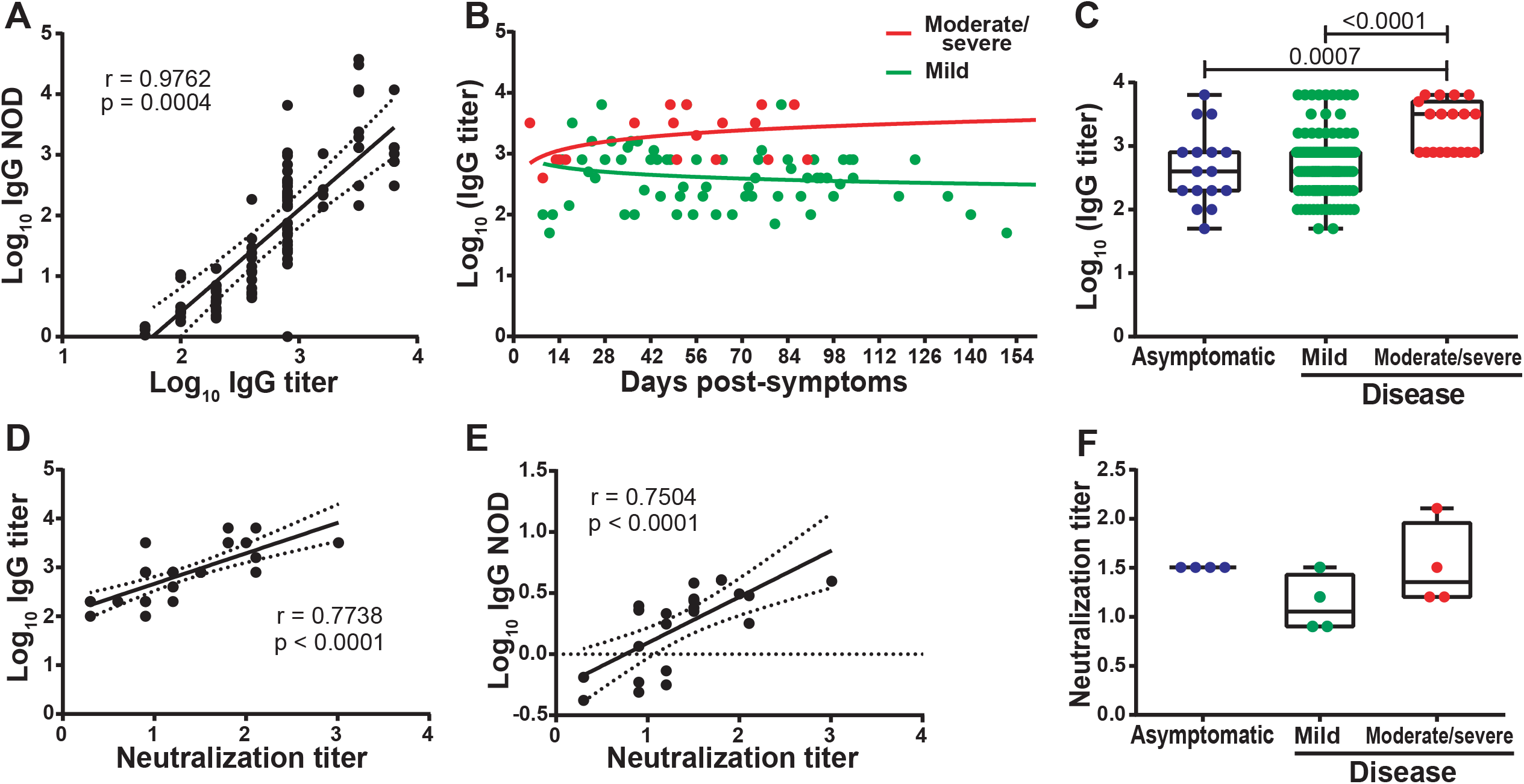
Relationship between optical density, titer, and neutralizing capacity of anti-SARS-CoV-2 IgG antibodies. **A)** Correlation analysis between Log10 of normalized optical density (NOD) and Log10 of IgG titers. **B)** Log10 of IgG titers corresponding to individuals who experienced moderate/severe (red dots) or mild disease (green dots) were plotted by day post-symptoms onset. **C)** Log10 of IgG titers from asymptomatic, and symptomatic individuals. **D)** Correlation analysis between Log10 IgG titers and IgG neutralizing titers and **E)** Log10 of NOD IgG values versus and IgG neutralizing titers. **F)** IgG neutralizing titers from asymptomatic, and symptomatic individuals. Individual measures, median and 25th and 75th percentiles are shown. Correlation studies were performed by using the Spearman rank test. In B, longitudinal data were studied by multiple linear regression, with disease severity and days post symptoms as independent predictors. Statistical comparisons between groups in C and F, were made by using Kruskal-Wallis test followed by Dunn’s post-test.

A subset of samples (N=34) were then tested for their capacity to neutralize WT SARS-CoV-2 virus. Neutralization titers showed moderate to strong direct correlations with Log_10_ (IgG titers) (Spearman’s correlation r=0.7738, p<0,0001; Figure 3D) and Log_10_ (IgG NOD) (Spearman’s correlation r=0.7504, p<0,0001; Figure 3E). The data support the use of anti-spike and anti-RBD IgG levels as a surrogate of IgG titers and, most important, SARS-CoV-2 neutralization titers.

In order to provide a deeper insight into the characteristics of the antibody response in asymptomatic donors and in those who experienced a mild or moderate/severe disease, 4 donors from each group with equal IgG titer (i.e. 800) were selected and used to evaluate viral neutralization capacity. Results indicated that, within a given IgG titer, asymptomatic donors develop neutralizing antibodies to the same extent that severe disease donors (Figure 3F).

### Complementary analysis in donors with discordant diagnostic results (positive SARS-CoV-2 PCR and non-detectable antibodies) and highly exposed SARS-CoV-2 seronegative contacts (ES)

In order to rule out the possibility that PCR^+^/ab^neg^ donors as well as donors consigned as ES donors had antibody responses but not detected by the COVIDAR ELISA, we used two additional assays in selected samples to solve discordant diagnostic results, to discard false negative results and to properly categorize donors within the cohort. First, we used an in-house IF assay to detect SARS-CoV-2 specific IgG. Out of 65 selected samples from PCR^+^/ab^neg^ donors, 46 were negative in the IFA, 7 were indeterminate (did not reach positive criteria), and 2 resulted in an unspecific reaction (signal was observed both in infected cells and in the uninfected control) (Figure 4A). The 7 samples with indeterminate result were also tested using the Abbot Alinity assay and resulted IgG negative. Thus, in all these 55 cases, the COVIAR result was confirmed and donors remained categorized as donors with molecular diagnostic of SARS-CoV-2 infection but no evidence of IgG seroconversion. On the other hand, 10 samples had detectable SARS-CoV-2-specific IgG by IFA. Out of these, 3 were also positive in the Abbot Alinity assay, 6 were negative and 1 was not tested. These 10 cases were re-categorized as donors with signs of seroconvertion to SARS-CoV-2-specific IgG based on IFA (Figure 4A). In the case of one of these donors (a donor whose initial sample was obtained 35 days after symptom onset and whose results were COVIDAR not detectable, IFA positive and Alinity not detectable), we could obtain a follow-up sample (at day 90 after symptom onset) that now gave a detectable result in the COVIDAR kit, confirming that the IFA result was correct. Probably, the broader IFA antigenic configuration helped detect IgG antibodies earlier in this case.

**Figure 4:**
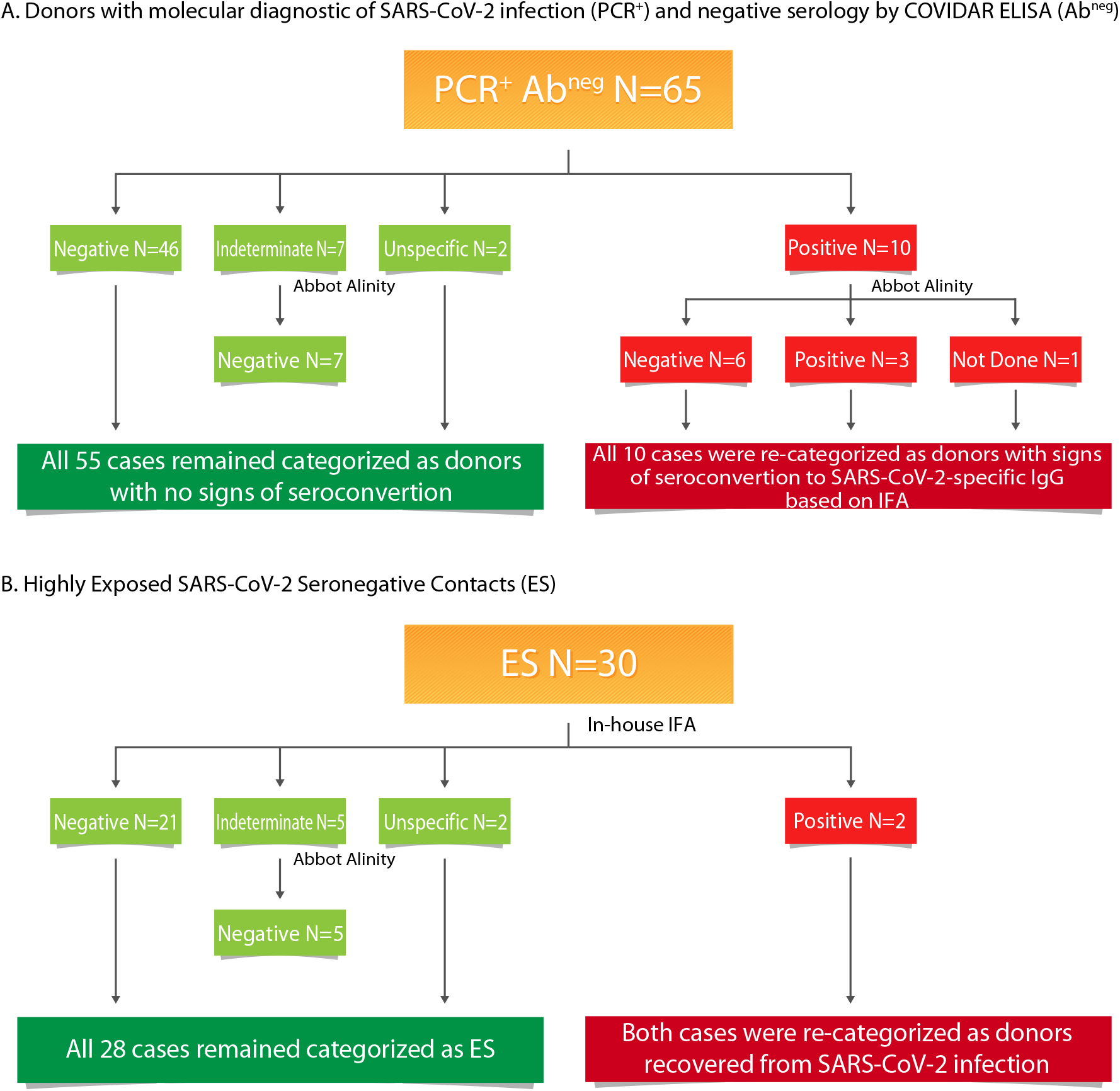
Flowchart indicating serology results obtained using different methodologies to evaluate SARS-CoV-2-specific IgG. **A)** Analysis in a subgroup of donors with molecular diagnostic of SARS-CoV-2 infection (PCR+) and undetected SARS-CoV-2-specific IgG by the COVIDAR kit (Ab^neg^). B) Analysis in a subgroup of suspected highly exposed uninfected donors.

On the other hand, 30 arbitrary selected EU were also tested by IFA. Twenty-one tested negative, 5 were indeterminate, 2 resulted positive and in other 2 unspecific reactions were observed. Out of the 5 indeterminate, none had a detectable result at the Abbot Alinity assay. Thus, 28 EU remained categorized as such. Contrary, 2 donors had detectable SARS-CoV-2-specific IgG by IFA, so they were recategorized as infected recovered donors (Figure 4B).

Additionally, the biobank had a set of 4 particular donors. None of them had molecular diagnostic done, all were antibody negative by COVIDAR but had positive antibody results by other commercially available ELISA kits that had been performed in other settings (private laboratories). One of them was positive by IFA and Abbot Alinity, one was indeterminate by IFA and positive by Abbot Alinity, one was positive by IFA and negative by Abbot Alinity and one was negative by IFA (Abbot Alinity was not performed in this case). Moreover, plasma from these donors were evaluated for neutralization activity. Two of them showed clear inhibition of viral replication with neutralization titers of 8, and >32, respectively, confirming the presence of SARS-CoV-2-specific antibodies in these samples. Thus, three donors were categorized as confirmed SARS-CoV-2 cases based on the criteria of having at least one detectable serological test. The fourth was also categorized as confirmed SARS-CoV-2 case based on the external serology result and an incident was added to its records in order to denote this issue.

Overall, the use of multiple assays with different configurations helped rule out the discordant diagnostic results, allowed to confirm the infection in these cases, and permitted the re-categorization of at least 16 cases. It is worth noting that these results were accounted for the final segregation of donors into the corresponding category presented at the initial section of this work.

### Changes in SARS-CoV-2-specific IgG titers after plasma donation

A group of donors belonging to the collection also donated plasma for treatment at different health institutions from Buenos Aires. Six of them returned at different times after plasma donation, so we were able to measure IgG titers before and after donation (plasma donors). For comparison, IgG titers were also evaluated in longitudinal samples from individuals who did not donate plasma (plasma non-donors) (Figure S2). Figure 5A shows the IgG titers at different time-points evaluated in the 6 plasma donors and in 5 plasma non-donors. At first sight, it can be observed that plasma donors show a steeper decay in IgG titer than plasma non-donors. It is also true if we compare these curves with the titer curve of the whole group (Figure 2B). When IgG antibody titers were analyzed in function of time elapsed from day of symptoms onset using a repeated measures lineal regression model, plasma donors showed indeed a stepper decay than plasma non-donors (p=0.0006, Figure 5B). Although the number of samples in this analysis is low small, this observation deserves further analysis as administration of convalescent plasma is one possible treatment available to prevent disease worsening in at-risk individuals [13]. It is important to highlight that SARS CoV-2 specific cellular response did not change over time in these individuals when Spike- and RBD-specific IFN-*γ* secreting cells were evaluated by ELISPOT (Figure S3). Thus, it is important to identify factors affecting antibody levels in plasma units to be used for treatment.

**Figure 5:**
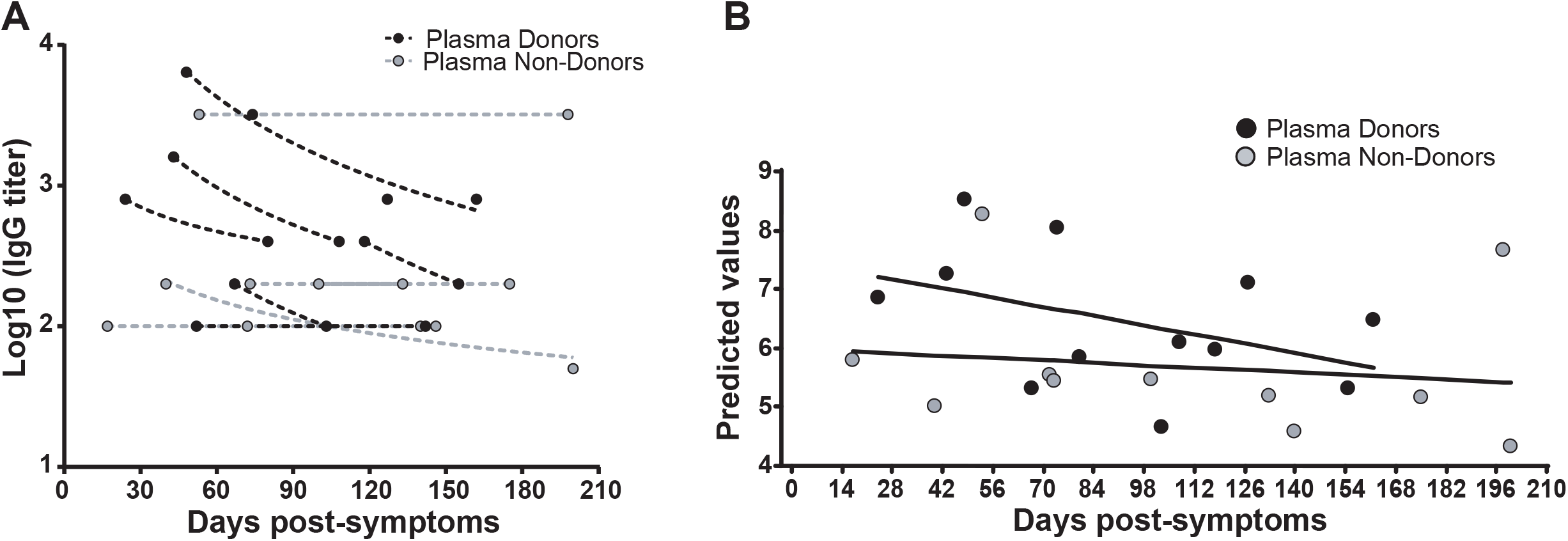
Fluctuations in IgG titers due to convalescent plasma donation. **A)** Antibody titers from samples obtained both before and after convalescent plasma donation were quantified, and plotted by days post-symptom onset (black dots, plasma-donors). Time-matched samples from plasma non-donors were analyzed as controls (grey dots, plasma non-donors). **B)** A repeated measures lineal regression analysis was applied to model data. Plasma donation, days since the onset of symptoms, as well as the interaction between them were set as fixed predictors, and subjects as random effects. Fitted lines and linear prediction equations for Log10 IgG titers in plasma are shown.

## CONCLUSIONS

Biobanks are key entities to accelerate basic, preclinical, translational, and clinical research. As SARS-CoV-2 spread worldwide, there was, and still is, an urgent need to learn more about this virus, to understand and treat the spectrum of clinical manifestations it produces, as well as to stop its spreading in the human population. As a result, the demand from the research community for samples from infected or recovered patients, as well as its related clinical data, has increased dramatically. In this context, the BBEI rapidly rearranged its procedures in order to cope with this need and started to collect, process and store COVID-19 samples and data, very early after SARS-CoV-2 landing in Argentina. Concomitantly, an in-depth clinical, immune and genetic characterization of this collection is being performed. This adds an extra value to the collection since all the data generated will be available upon request which represents a tool to accelerate research and development projects. In this initial report (encompassing data from 825 donors), the dynamics, magnitude and quality of humoral response within a period of 5 months after symptom onset is described, and also changes in IgG dynamics in recovered participants that donated plasma for therapeutic purposes.

In the first place, it was observed that there exists an extraordinary heterogeneity in the level of SARS-CoV-2 spike- and RBD-specific IgM and IgG antibodies among individuals. As others reported, specific IgM and IgG appeared simultaneously,. IgM levels did not achieve a plateau, instead they showed a slight decrease. Nevertheless, a significant proportion of donors had detectable specific IgM up to 100 days after symptom onset. This limits IgM utility as a marker of acute or “active” infection and also opens the question regarding its usefulness in identifying possible reinfections since a detectable result could be due to carry-over from the first episode. Contrary, specific IgG seems to be more stable over time with a high positivity rate even in samples obtained after 120 days following symptom onset. In a breakthrough report, Ibarrondo et al described that SARS-CoV-2 IgG antibodies decayed early and rapidly after symptom onset in persons with mild disease [14]. Other studies confirmed that indeed there is a decay in IgG levels but at a slower rate and at later times than that reported by Ibarrondo et al [11, 15-21]. Although our understanding in the antibody dynamics is still incomplete, specific IgG levels appear to be maintained during 4-5 months, or even longer, if we consider our data from plasma non-donors, presented in figure 5A. After 5 months, the magnitude of the humoral response may wane, however, detectability rate remains high. The longevity of the response might be influenced by a number of factors such as age, number of symptoms and the severity of the disease (Figure 2), in agreement with previous reports. Additionally, the magnitude of the peak response and other yet-to-be-defined factors might shape this response. On the other hand, it is still unclear why some asymptomatic persons develop robust humoral responses while others do not. We are starting to understand that SARS-CoV-2-specific memory B cells are maintained by 6 months post-infections and continue to evolve [16, 18, 22]. Thus, despite antibody declining, this memory response could confer long-term protection to subsequent exposures, disease progression after a re-infection and it could determine vaccine efficacy. Certainly, more data is needed to complete the picture.

A relevant finding in our cohort is the high percentage of individuals who had molecular diagnostic of infection but no evidence of seroconversion after 21 days of symptoms onset. The analysis with different methodologies to detect SARS-CoV-2-specific antibodies, within this subset of donors, and the study of follow-up samples, allowed to evidence, at least in a minor proportion of these donors, the presence of SARS-CoV-2 specific antibodies. Several reports in the literature indicate that combined antibody detection through multiple assays with different specificities helps to increase the sensitivity and specificity in serological diagnostics [23-25]. Still, specific antibodies remained undetectable in a considerable high proportion of donors (15.2% of the whole cohort), compared to other reports that describe only 5-10% or even less [26-28]. One possible reason to explain this discrepancy could be associated with false positive results of the molecular diagnostic, which was proven not so infrequent in the context of mass testing [29, 30]. Second, the highest rates of antibody detectability are shown in studies involving persons who were or had been hospitalized while our study might be biased due to the inclusion of a number of asymptomatic donors, mainly represented by health workers screened during routine surveillance at their hospitals. In turn, this leads to a third hypothesis, which is the possibility of SARS-CoV-2 carriage without infection, as has already been described for other respiratory viruses (including common coronaviruses) [31] and that can even be overcome with a protective innate immune response leading to an abortive infection [32, 33]. Finally, the possibility that these donors developed a T-cell response without achieving a detectable antibody production, as was described earlier for SARS-CoV-2 and other viruses such as Hepatitis C virus, cannot be ruled out [34, 35]. In fact, it has been observed that T-cell responses could last and be robust in the absence of anti-SARS CoV-2 antibody detection in close contacts from COVID-19 patients with mild symptoms from France [34]. In that report, authors stated that COVID-19 contacts might have developed very low levels of antibodies, which could not be detected by the assays used in that study. More recently, Schwarzkopf et al reported that 17% of convalescent potential plasma donors participating in their study had borderline or negative antibody testing while most of them had T-cell immunity against SARS-CoV-2 [36]. Evaluation of SARS-CoV-2-specific T-cell responses in our cohort is ongoing. Besides antibody binding capacity, determining neutralization capacity is key to understand the role of humoral response in the natural course of the disease. There is wide consensus that a robust neutralizing antibody response rise early following infection and that this response is mostly determined by the magnitude of anti-spike and anti-RBD IgGs [11, 17, 20, 37-40]. Measuring neutralization activity against WT SARS-CoV-2 can be labor intensive and is restricted to institutions with BSL-3 facilities and properly trained personnel. Alternative protocols have been developed using pseudotyped viruses but still infrastructure limitations apply. Here, we confirm that the level of S-specific and RBD-specific IgG antibodies measured by the COVIDAR kit (either as IgG NOD or IgG end-point titer) can function as surrogate markers of neuralization activity against WT SARS-CoV-2 (Figure 3E). This has also been previously demonstrated in an independent work using SARS-CoV-2 pseudotyped VSV particles [11].

The use of plasma from persons recovered from COVID-19 was one of the first treatment administered. There is growing evidence that early infusion is effective in preventing disease worsening but its success relies on the use of plasma units with high titers of SARS-CoV-2-specific antibodies and early administration [13, 41-44]. Limitations for this treatment include donor eligibility (exclusion criteria apply), willingness of recovered persons to donate (probably more than one time), early administration, logistic issues, among others [45, 46]. Other antibody-based therapies have been developed to replace the use of convalescent plasma, such as monoclonal antibodies and hyperimmune equine serum, but at higher costs. Thus, it is worth investing efforts to improve the scope of this treatment. Here, we observed that SARS-CoV-2 IgG titers decay rate would be more rapid after plasma donation. Even though it has not been previously described, this observation was not completely unexpected. Plasmapheresis has been used to treat humoral rejection in kidney transplant by reducing antibody levels [47]. Although our observation still needs to be confirmed in a larger cohort and in a specially designed work, it still deserves attention since it might be a factor to be mitigated in order to maximize the benefit in plasma recipients. In this regard, concern has already been raised highlighting the need of antibody measurement at the time of plasmapheresis based on the spontaneous decay in IgG titer [48]. Moreover, optimum timing from symptom onset to plasma donation has been proposed [49, 50]. Perreault et al evaluated anti-RBD IgG in plasma donors who donated multiple times [51]. They found that the anti-RBD antibody levels waned over time as a consequence of time and not of the number of donations. However, they did not have a non-donor group to compare the declining rate as in our study. Thus, antibody levels should be unavoidable measured before each plasmapheresis, even for individuals who donate multiple times within a few days, in order to guarantee the quality, in terms of specific IgG titers, of the plasma to be used. At the individual level, it is unlikely that protective immunity could be affected in sequential plasma donors. So far, there is no evidence that plasma donors are more susceptible to re-exposures. Indeed, the magnitude of SARS-CoV-2-specific T-cell responses seems to be unchanged pre- and pos-donation thus protection would be conferred by cellular memory responses.

SARS-CoV-2 pandemic has driven the world to an unprecedented crisis, not only in terms of public health but also social, financial and productive. However, it has highlighted the role of basic and translational science in the society. In this context, biobanks have gained visibility within research community but also in the community as a whole, mostly driven by the ability to widely and immediately share samples and data from COVID-19 patients with warranted quality thus accelerating research, and by engaging community to promote sample donation. Moreover, the BBEI-COVID19 collection served a dual role in this emergency situation. First, it accomplished the main aim of creating a tool to boost researchers and developers transferring samples upon request for approved projects. By the date this manuscript was submitted, 1030 donors were included in the collection and 927 biological specimens were transferred to 9 biomedical research projects, while other 335 samples have been requested, and are awaiting for committee approval. At the same time, it generated highly needed local data of extreme importance to understand and frame the regional dynamics of the infection. As we can expect that this pandemic will continue to be of paramount concern for months to come, work at biobanks will continue filling in the gaps of existing knowledge as well as dissecting future challenges such as the effect of vaccines on the pandemic dynamics or the potential of reinfections.

## Data Availability

All the information included in the manuscript is available upon reasonable request and prior approval of the biobank directory board.

## ACKNOWLEDMENTS

We profoundly thank COVIDAR working group for kits supply, technical advice and data interpretation and discussion. We particularly thank Andrea Gamarnik for insightful inputs into the manuscript. COVIDAR working group: Diego OJEDA, María Mora GONZALES LEDESMA, Guadalupe S. COSTA NAVARRO, Horacio M. PALLARES, Lautaro N. SANCHEZ, Sergio M. VILLORDO, Diego E. ALVAREZ, Julio J. CARAMELO, Jorge J. CARRADORI, Marcelo J. YANOVSKI y Andrea V. GAMARNIK (Fundación Instituto Leloir-CONICET, Buenos Aires, Argentina).

Authors specially acknowledge to BBEI donors for agreeing to collaborate with the biobank and to provide blood samples.

This work was supported by a grant from the *Agencia Nacional de Promoción de la Investigación, el Desarrollo Tecnológico y la Innovación* (Agencia I+D+i) from Argentina through an extraordinary funding opportunity to improve the national response to COVID19 (Proyecto COVID N° 11, IP 285). The funders had no role in study design, data collection and interpretation, or the decision to submit the manuscript for publication. We also thank Mr. Fernando Montesano for his invaluable help with donor’s admission and data entry.

## Supplementary Figures

**Figure S1: A**. Distribution of donors with past or present confirmed SARS-CoV-2 infection per group (Total N=735). Infection was confirmed either by molecular diagnostic, serology or both. **B**. SARS-CoV-2-specific IgM and IgG positivity rate per group. Antibodies were evaluated in plama samples by ELISA using the COVIDAR kit. AS: acute severe, AM: acute mild, ECS: early convalescent severe, ECM: early convalescent mild, LCS: late convalescent severe, LCM: late convalescent mild.

**Figure S2:** Characteristic of plasma donors and non-donors included in Figure 5.

**Figure S3:** Spike- and RBD-specific IFN-γ-secreting cells measured by ELISPOT in samples obtained from plasma donors pre- and pos-donation.

