## Supplemental Figure 1 for "Dynamics of SARS-CoV-2-specific antibodies during and after COVID19: Lessons from a biobank in Argentina"

### Supplementary Figure 1.

**A.** Distribution of donors with past or present confirmed SARS-CoV-2 infection per group (Total N=735). Infection was confirmed either by molecular diagnostic, serology or both. **B.** SARS-CoV-2-specific IgM and IgG positivity rate per group. Antibodies were evaluated in plama samples by ELISA using the COVIDAR kit. AS: acute severe, AM: acute mild, ECS: early convalescent severe, ECM: early convalescent mild, LCS: late convalescent severe, LCM: late convalescent mild.

**A.**

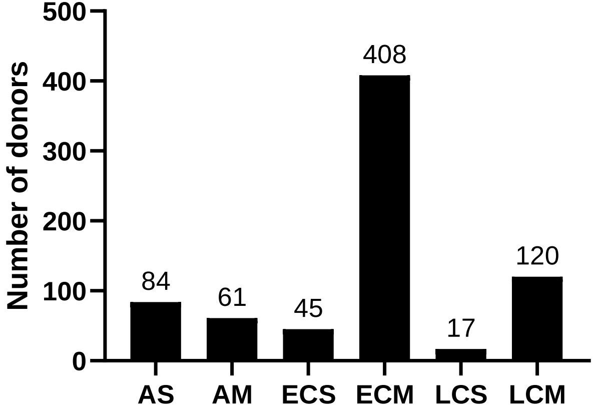


**B.**

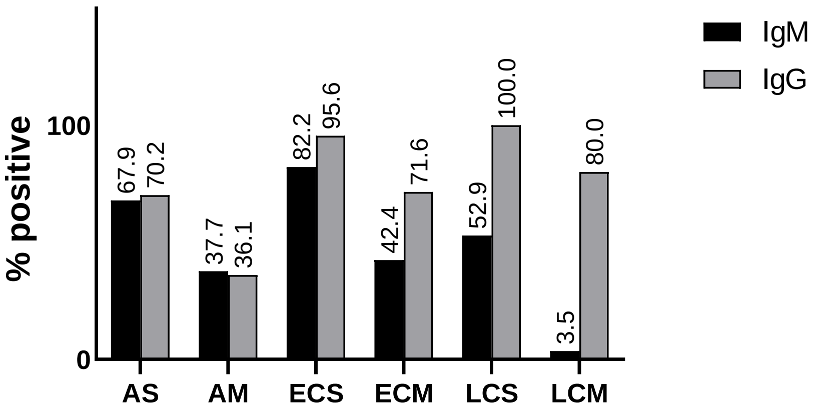
