## Supplemental Figure 2 for "Dynamics of SARS-CoV-2-specific antibodies during and after COVID19: Lessons from a biobank in Argentina"

Supplementary figure 2: Characteristic of plasma donors and non-donors included in Figure 5

| Group | Donor ID* | Gender | Age range | Disease Course | Time from symptom onset to 1st sample (days) | Time between 1st and 2nd sample (days) | Time between 1st and 3rd sample (days) | Number of plasmapheresis | IgG Titer Sample 1 | IgG Titer Sample 2 | IgG Titer Sample 3 |
|---|---|---|---|---|---|---|---|---|---|---|---|
| Plasma Donor | PD1 | Male | 60-69 | Severe | 48 | 79 | 114 | 3 | 3200 | 800 | 800 |
| Plasma Donor | PD2 | Female | 20-29 | Mild | 52 | 90 | | 1 | 100 | 100 | |
| Plasma Donor | PD3 | Female | 30-39 | Mild | 43 | 65 | | 1 | 1600 | 400 | |
| Plasma Donor | PD4 | Male | 40-49 | Mild | 67 | 36 | | 2 | 200 | 100 | |
| Plasma Donor | PD5 | Female | 20-29 | Asymptomatic | - | 59 | | 1 | 800 | 400 | |
| Plasma Donor | PD6 | Male | 40-49 | Mild | 118 | 37 | | 1 | 400 | 200 | |
| Plasma Non-Donor | PND1 | Male | 40-49 | Mild | 40 | 100 | 160 | 0 | 200 | 100 | 50 |
| Plasma Non-Donor | PND2 | Male | 60-69 | Severe | 48 | 26 | | 0 | 6400 | 3200 | |
| Plasma Non-Donor | PND3 | Male | 20-29 | Mild | 73 | 60 | | 0 | 200 | 200 | |
| Plasma Non-Donor | PND4 | Male | 40-49 | Asymptomatic | 53** | 145 | | 0 | 3200 | 3200 | |
| Plasma Non-Donor | PND5 | Male | 30-39 | Mild | 17 | 74 | 129 | 0 | 100 | 100 | 100 |
| Plasma Non-Donor | PND6 | Male | 30-39 | Mild | 100 | 75 | | 0 | 200 | 200 | |

*Donor IDs shown in this table are arbitrary and have no association with the biobank code.

**Time was estimated taking into account the date of diagnostic of a household contact.
