## Supplemental Figure 3 for "Dynamics of SARS-CoV-2-specific antibodies during and after COVID19: Lessons from a biobank in Argentina"

Supplementary Figure 3.

IFN-γ-secreting cells were detected using enzyme-linked immunospot (ELISPOT) assays conducted as described previously. Briefly, 200.000 PBMC were plated on sterile 96-well plates (MultiScreen IP plates; Millipore) in the presence of Spike or RBD protein (10 ug/mL). Plates were developed using biotinylated anti-human IFN-γ monoclonal antibody, streptavidin peroxidase complex, and AEC (3-amino-9-ethylcarbazole) substrate reagent set (BD Biosciences). Plates were scanned on an ImmunoSpot reader (Cellular Technology Ltd.). Specific spots were counted using the ImmunoSpot software. Results were expressed as spot forming units (SFU)/106 PBMC. Data was analyzed using Wilcoxon test for pair samples with non-parametric distribution.

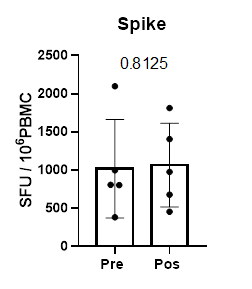

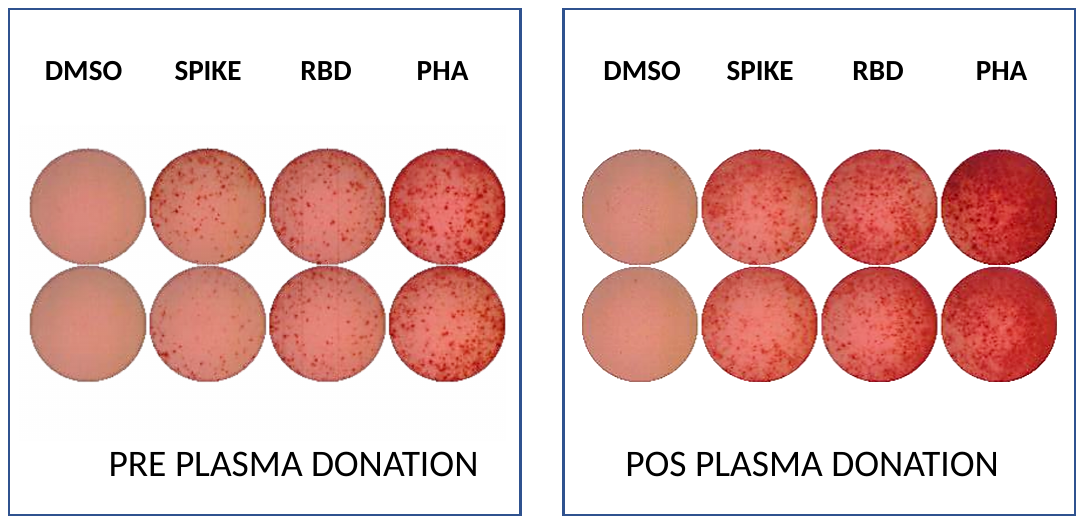

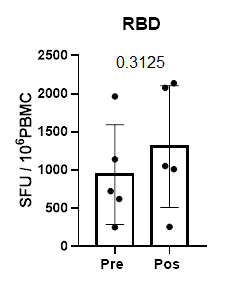
